# Predictive Value of Neutrophil-to-Lymphocyte and Platelet-to-Lymphocyte Ratios for Distant Metastasis in Vietnamese Gastric Cancer Patients

**DOI:** 10.64898/2026.01.05.25342950

**Authors:** N. Tran, L. Tran Dang Ngoc, Q.H. Nguyen

## Abstract

**Background:** Gastric cancer (GC) remains a leading cause of global cancer mortality, with a high prevalence of distant metastasis at diagnosis in Vietnam. Systemic inflammatory markers are emerging as potential tools for risk assessment, yet their independent predictive value in the Vietnamese population requires further clarification.

**Objective:** To evaluate the predictive value of the Neutrophil-to-Lymphocyte Ratio (NLR) and Platelet-to-Lymphocyte Ratio (PLR) as cost-effective biomarkers for distant metastasis in Vietnamese GC patients.

**Design:** A single-center, retrospective cohort study of 114 GC patients treated at the Oncology Hospital of Ho Chi Minh City between January and June 2023.

**Methods:** Patients were categorized into metastatic (Stage IV) and non-metastatic groups per AJCC 8th edition criteria. Pretreatment NLR and PLR were calculated from peripheral blood counts. The optimal cutoffs were determined using receiver operating characteristic (ROC) curve analysis. DeLong’s test was employed to compare the diagnostic efficacy of the clinical model versus the combined clinical-hematological model. Univariate and multivariate binary logistic regression analyses were performed to identify independent predictors of distant metastasis.

**Results:** Metastasis was present in 44.7% of cases. Optimal cutoffs of 2.0 for NLR and 181.5 for PLR were identified. Both markers were significantly higher in the metastatic group (p<0.01) and positively correlated with disease progression. A combined model integrating clinical factors with NLR and PLR demonstrated superior diagnostic accuracy compared to clinical factors alone (AUC: 0.766 vs 0.619, p=0.0036). In multivariable analysis, multiple tumor location and high PLR remained independent prognostic factors for distant metastasis.

**Conclusion:** Elevated NLR and PLR are significantly associated with distant metastasis in GC. Integrating these accessible, minimally invasive markers with clinical data enhances predictive accuracy, providing a practical tool for risk stratification in resource-limited settings.

**Plain Language Summary:** *The Challenge:* Gastric cancer (stomach cancer) is a leading cause of cancer-related deaths worldwide. In Vietnam, many patients are diagnosed only after the cancer has already spread to distant parts of the body (metastasis). When cancer spreads, it becomes much harder to treat. Doctors need affordable and reliable ways to predict which patients are at a higher risk of metastasis to improve how they manage the disease.

*The Study:* Researchers studied 114 gastric cancer patients at the Ho Chi Minh City Oncology Hospital. They looked at two specific markers found in routine, inexpensive blood tests: the Neutrophil-to-Lymphocyte Ratio (NLR) and the Platelet-to-Lymphocyte Ratio (PLR). These markers measure the balance of different white blood cells and platelets, which partially reflect the body’s “inflammation” levels in response to a tumor.

*The Findings:* The study found that patients with advanced, metastatic cancer had significantly higher NLR and PLR levels compared to those in earlier stages. By combining these blood markers with other clinical information—such as the patient’s and tumor’s data —the researchers created a predictive model. This combined model was much more accurate at identifying patients with distant metastasis than using clinical information alone.

*The Impact:* Because these blood tests are simple, low-cost, and already widely available in hospitals, they offer a practical way for doctors to monitor cancer progression. Using the NLR and PLR ratios can help healthcare providers in Vietnam and elsewhere better identify high-risk patients and personalize their treatment plans more effectively.

## Introduction

Gastric cancer (GC) is one of the most common cancers in the world, ranking fifth in incidence and fourth in mortality among all cancer types. In Vietnam, GC is among the five most prevalent cancers in both sexes, accounting for 9.8% of all cancer diagnoses.^[1]^

Despite significant advances in detection and disease management through screening recommendations and *Helicobacter pylori* eradication strategies, the survival prognosis for GC remains poor. This is largely due to the fact that most cases present with distant metastases at the time of diagnosis,^[2^, ^3^] underscoring the need for thorough investigation of distant metastasis at initial consultation.

Several clinical and pathological features such as age, sex, ethnicity, location of the primary tumor, depth of invasion, lesion size, and histological differentiation have been reported as risk factors for distant metastasis in GC.^[4]^ Tumor markers including CEA, CA 19-9, and CA 72-4 have primarily been reserved for longitudinal monitoring rather than initial screening, as their diagnostic performance varies significantly across different clinical reports.^[5–8]^ Regarding imaging modalities, PET/CT is an effective strategy for screening distant metastases; however, its high cost and limited accessibility hinder its widespread use.^[9]^

It has been reported that inflammation is closely linked to cancer pathogenesis, playing an important role in disease progression and determining patients’ clinical outcomes. The activity of inflammatory mediators, cytokines, and immune cells within the tumor microenvironment (TME) have been reported to contribute to the initiation, development, progression, and distant metastasis of GC.^[10]^

Various systemic inflammatory-immune markers have been tested over the past two decades to serve as potential biomarkers for disease surveillance and prognosis. Notably, the cellular components of leukocytes involved in inflammation and antitumor immune responses are currently promising indicators of disease status.

Neutrophils are responsible for the initial immune response against invading pathogens through mechanisms such as chemotaxis, phagocytosis, and release of reactive oxygen species (ROS).^[11]^ They also play a crucial regulatory role in adaptive immune responses by secreting a range of pro-inflammatory cytokines and chemokines to enhance the recruitment, activation, and regulation of other immune cells. ^[12]^ Lymphocytes are critical for immune responses to eradicate malignant cells and the reduction of lymphocytes is often associated with suboptimal antitumor immune response and disease progression.^[13^, ^14^] Besides, another compartment in complete blood count should be noticed is the platelet. Platelet aggregation can foster tumor growth by releasing angiogenic mediators within the tumor microvasculature.^[15]^ This cell type also enhances tumor cell adhesion and dissemination in the extracellular matrix, thereby promoting cancer cell proliferation and metastasis.^[16]^

Dynamic changes in peripheral blood cell populations, particularly the neutrophil-to-lymphocyte ratio (NLR) and platelet-to-lymphocyte ratio (PLR), have emerged as important biomarkers reflecting the systemic inflammatory response associated with cancer progression. Elevated NLR reflects a relative increase in neutrophils, which can secrete pro-inflammatory cytokines, growth factors, and proteases that facilitate tumor proliferation, invasion, and the establishment of a pre-metastatic niche. Concurrent lymphopenia indicates a compromised adaptive immune response, weakening cytotoxic T-cell activity and reducing immune surveillance against tumor cells.^[17^, ^18^] Similarly, an increased PLR suggests enhanced platelet activation, which supports tumor metastasis by protecting circulating tumor cells from immune destruction, promoting angiogenesis through the release of vascular endothelial growth factor (VEGF), and aiding in the adhesion of tumor cells to the endothelium at distant sites, collectively contributing to tumor aggressiveness and metastatic spread.^[19–22]^

These ratios have demonstrated prognostic value, correlating with disease severity and patient outcomes, across various solid tumors.^[23–27]^ However, their predictive utility specifically for distant metastasis in gastric cancer, particularly within the Vietnamese patient population, remains underexplored. Therefore, investigating the NLR and PLR in this context could provide valuable insights into their role as accessible, cost-effective markers for early identification of high-risk patients with distant metastasis, ultimately visualizing treatment strategies and improving clinical management of gastric cancer in Vietnam.

## Materials and Methods

### Study population and data collection

We collected the clinical data of gastric cancer patients at Ho Chi Minh City Oncology Hospital between January and June 2023. Patients were excluded if they met one of the following exclusion criteria: 1) concurrent or secondary malignant tumors at other sites. 2) Infection confirmed by laboratory tests. 3) Patients with systemic inflammatory conditions, history of autoimmune diseases, or currently undergoing anti-inflammatory treatment. 4) Patients diagnosed with hematological disorders or those who had received blood transfusions within the past three months. 5) Patients with organ failure (liver, kidney, heart, or other organs). 6) Patients with incomplete or unavailable pre-treatment medical records.

Ultimately, 114 patients with GC were enrolled in this study. We collected data on age, sex, ECOG performance status, BMI (Body Mass Index), tumor features (primary tumor location, macroscopic morphology of the lesion, and histological differentiation grade of tumor cells), and disease stage according to the American Joint Committee on Cancer (AJCC) 8^th^ edition. Pre-treatment laboratory results included total white blood cell (WBC, 10^9^ cells/L), absolute neutrophil count (ANC, 10^9^ cells/L), absolute lymphocyte count (ALC, 10^9^ cells/L), red blood cell count (RBC, 10^12^ cells/L), hemoglobin level (g/L), platelet count (PLT, 10^9^ cells/L), and other tumor markers such as CEA, CA19-9, and CA72-4, if available.

The NLR was calculated as the absolute neutrophil count divided by the absolute lymphocyte count. Similarly, PLR was calculated as the absolute platelet count divided by the absolute lymphocyte count.

Informed consent was obtained from all subjects and/or their legal guardian(s). All personal information of the patients included in the study was kept confidential and was used solely for research purposes. This study was approved by the hospital’s Ethics Committee of Ho Chi Minh City Oncology Hospital (Approval No. 292/BVUB-HĐĐĐ dated April 26, 2023).

### Statistical analysis

We first divided the patients into two groups: those with distant metastasis (stage IV) (n = 51) and those without distant metastasis (n = 63). Categorical variables are presented as percentages and compared between the two groups using the chi-square test (χ²) or Fisher’s exact test. The Kolmogorov-Smirnov test was used to test the normality of the continuous variables. Normally distributed continuous variables were expressed as mean ± standard deviation (SD) and compared between groups using Student’s t-test. In cases where continuous variables did not follow a normal distribution, variables were presented as median and interquartile range (IQR) and compared using the Wilcoxon-Mann-Whitney test.

The correlation between two continuous variables was determined using Pearson’s correlation test, whereas Spearman’s rank correlation test was used to assess correlations between non-parametric data.

The optimal cut-off values for NLR, PLR and predicted probability of models were estimated as the value with the highest using the Youden index (*J* = sensitivity + specificity – 1).

Receiver operating characteristic (ROC) analysis was used to evaluate the predictive performance of the binary logistic regression models for distant metastasis. Comparisons between two AUC (Area Under the Curve) values were performed using the DeLong test.

Univariate and multivariate logistic regression analyses were conducted to identify independent predictors of distant metastasis. Only variables significant in univariate analysis were included in the final multivariate model. The odds ratio (OR) estimated from the regression analysis is reported with a 95% confidence interval (95% CI).

A p-value < 0.05 was considered at significant for all comparisons, correlations, and regression analyses. All statistical analyses and visualizations were performed using the R software (version 4.3.0).

This study was completely checked according to the STROBE checklist,^[28]^ which was included in Table S1.

## Results

### Patient characteristics

For the entire cohort in our study, the mean age of the patients was 58 years (range 48–68 years). Males accounted for 62.28% of the GC cases, and the male-to-female ratio was 1.6:1. One-third of the patients (30.7%) had a BMI < 18.5.

Regarding tumor location, the antrum was the most common site (31.57%), followed by multiple locations (24.56%), and the corpus (20.17%). In terms of histopathological grade, more than half of the cases (57.89%) were diagnosed as poorly differentiated adenocarcinoma. The predominant macroscopic tumor type was exophytic mass (31.58%), followed by ulcerative (17.54%). Most patients were diagnosed at an advanced stage, with 31.57% classified as stage III and 44.73% as stage IV. Among the patients with distant metastases, peritoneal metastasis was the most frequent (18 cases, 22.5%), followed by mesenteric lymph node metastasis (15 cases, 18.75%), para-aortic lymph node metastasis (14 cases, 17.5%), and hepatic metastasis (12 cases, 15%). Most metastatic cases occurred at a single site (56.86%), whereas metastases affecting three or more sites were observed in 11.76% of cases (**Table 1**).

**Table 1.**
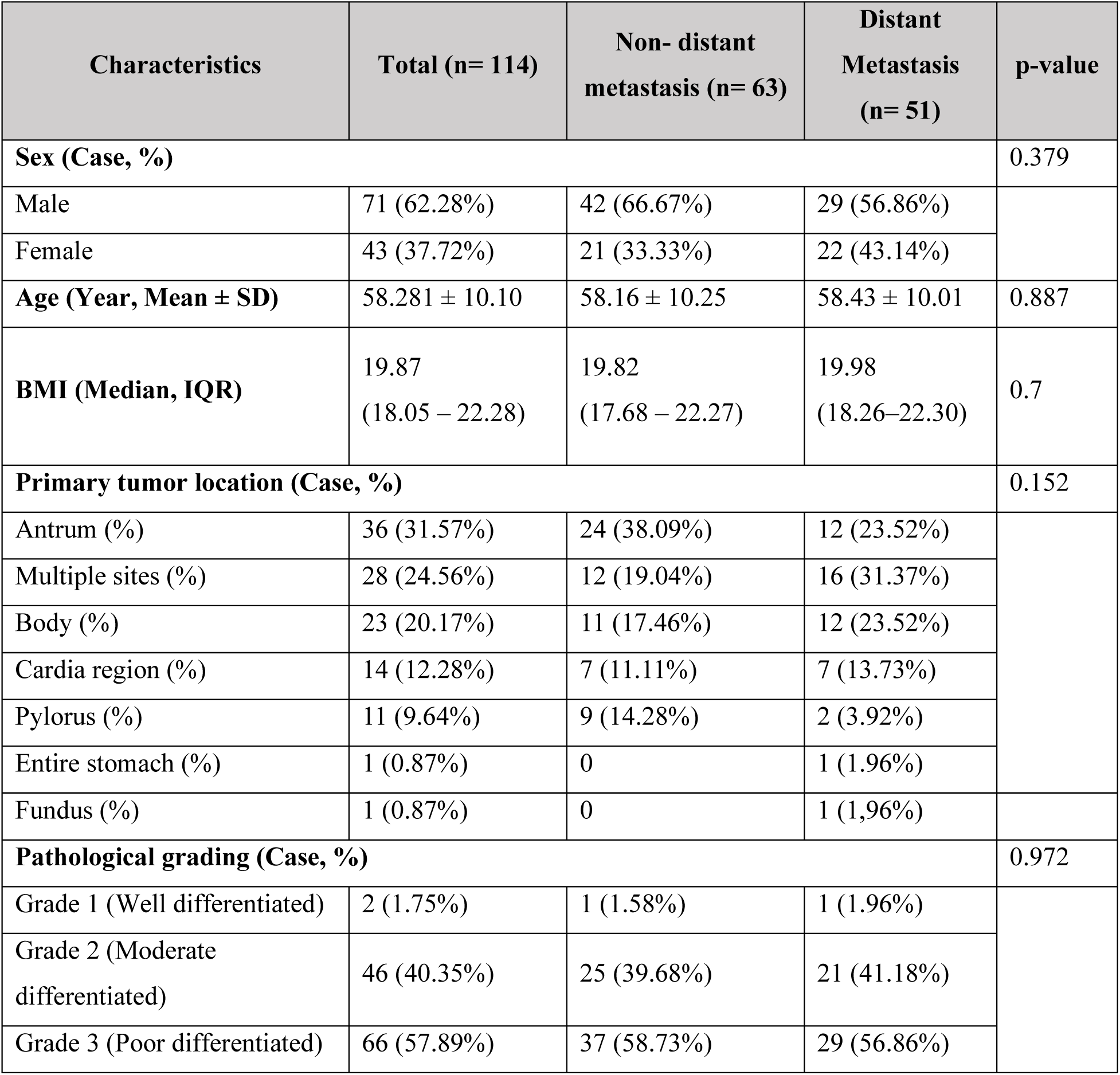
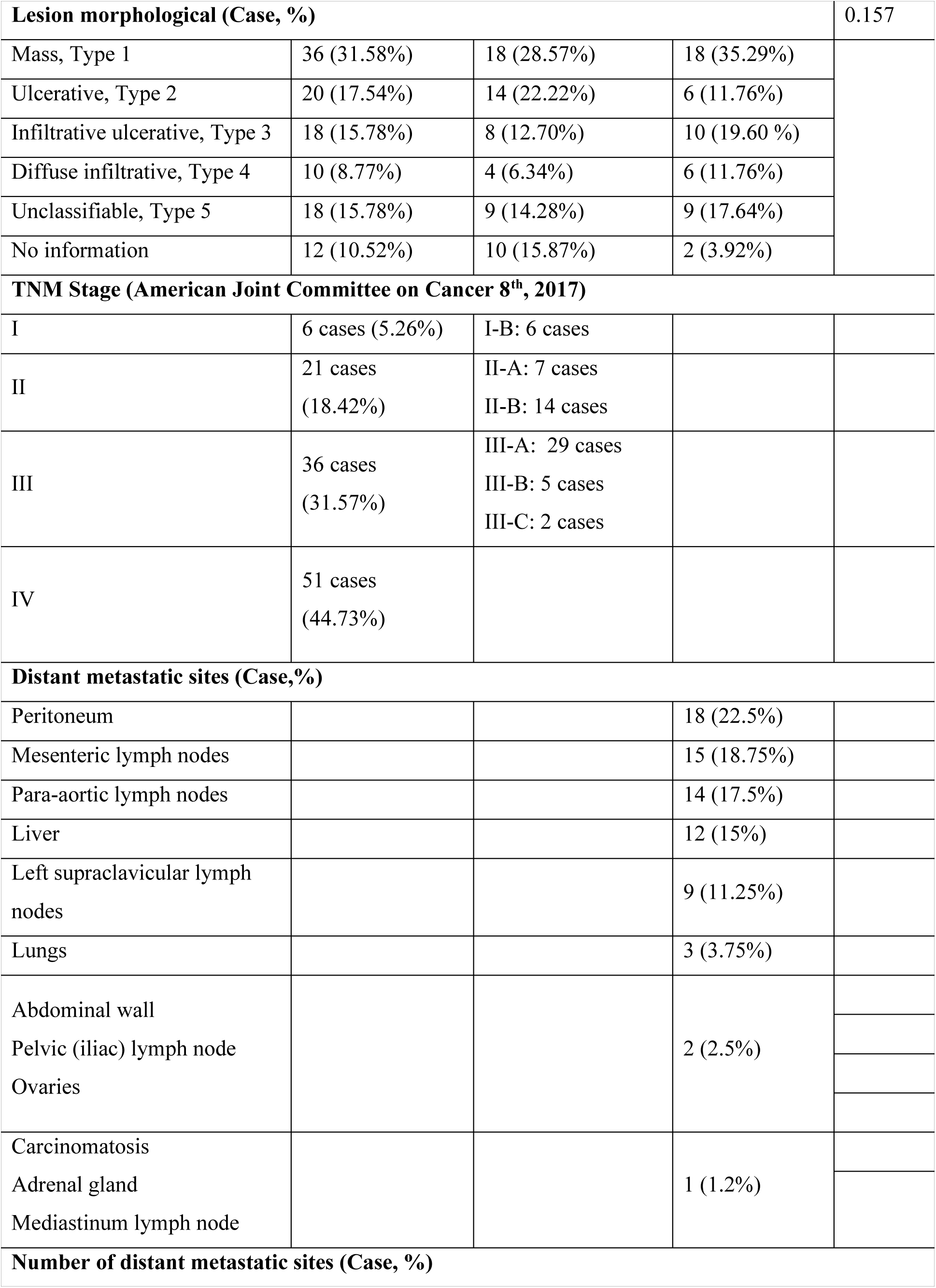
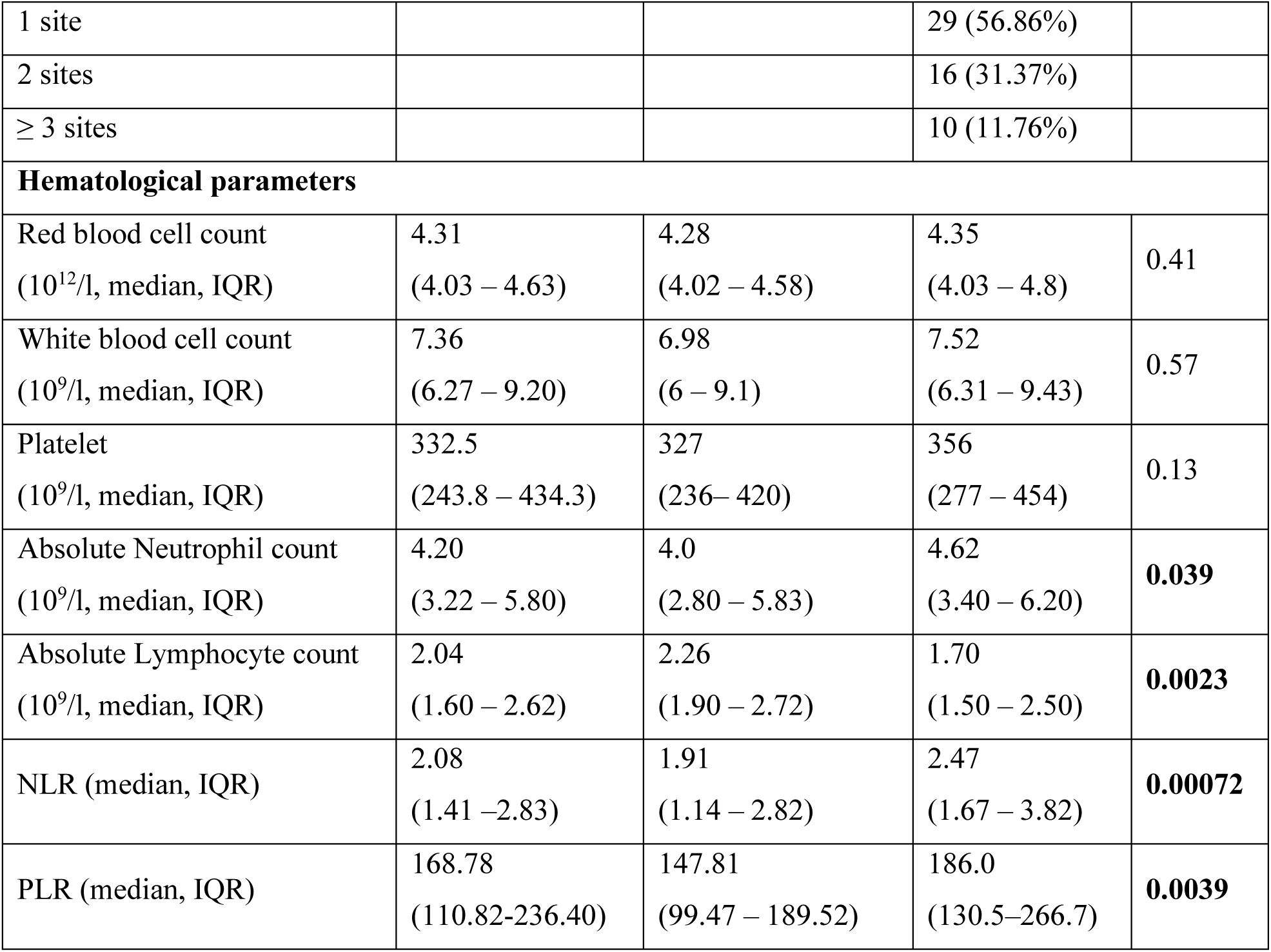
Baseline clinicopathological and hematological characteristics of patients. Continuous variables are presented as mean ± standard deviation (SD) for normally distributed data or median (interquartile range [IQR]) for non-normally distributed data. Categorical variables are reported as percentages (%). Abbreviations: BMI, Body Mass Index; TNM, tumor-node-metastasis; AJCC, American Joint Committee on Cancer; NLR, neutrophil-to-lymphocyte ratio; PLR, platelet-to-lymphocyte ratio; ANC, absolute neutrophil count; ALC, absolute lymphocyte count; RBC, red blood cell count; PLT, platelet count.

Hematological parameters showed that no significant differences were observed in red blood cell, platelet, or total white blood cell counts between two groups of patients with and without distant metastases (Wilcoxon test: red blood cells, *p* = 0.41; total white blood cells, *p* = 0.57; platelets, *p* = 0.13) (**Table 1**). However, we noticed that patients with distant metastases displayed a higher absolute neutrophil count compared to those without metastases, with median values of 4.62 × 10⁹ cells/L (IQR: 3.40–6.20) for metastasis group and 4.0 × 10⁹ cells/L (IQR: 2.80–5.83) for non-metastasis group, median difference 0.62 × 10⁹ cells/L (95% CI: 0.022–1.250), *p* = 0.039 (Wilcoxon test). Conversely, the absolute lymphocyte count was statistically lower in the metastatic group (1.70 × 10⁹ cells/L, IQR: 1.50–2.50) compared to the non-metastatic group (2.26 × 10⁹ cells/L, IQR: 1.90–2.72), median difference 0.56 × 10⁹ cells/L (95% CI: 0.095–0.594), *p* = 0.0023 (Wilcoxon test) (**Figure 1**).

**Figure 1.**
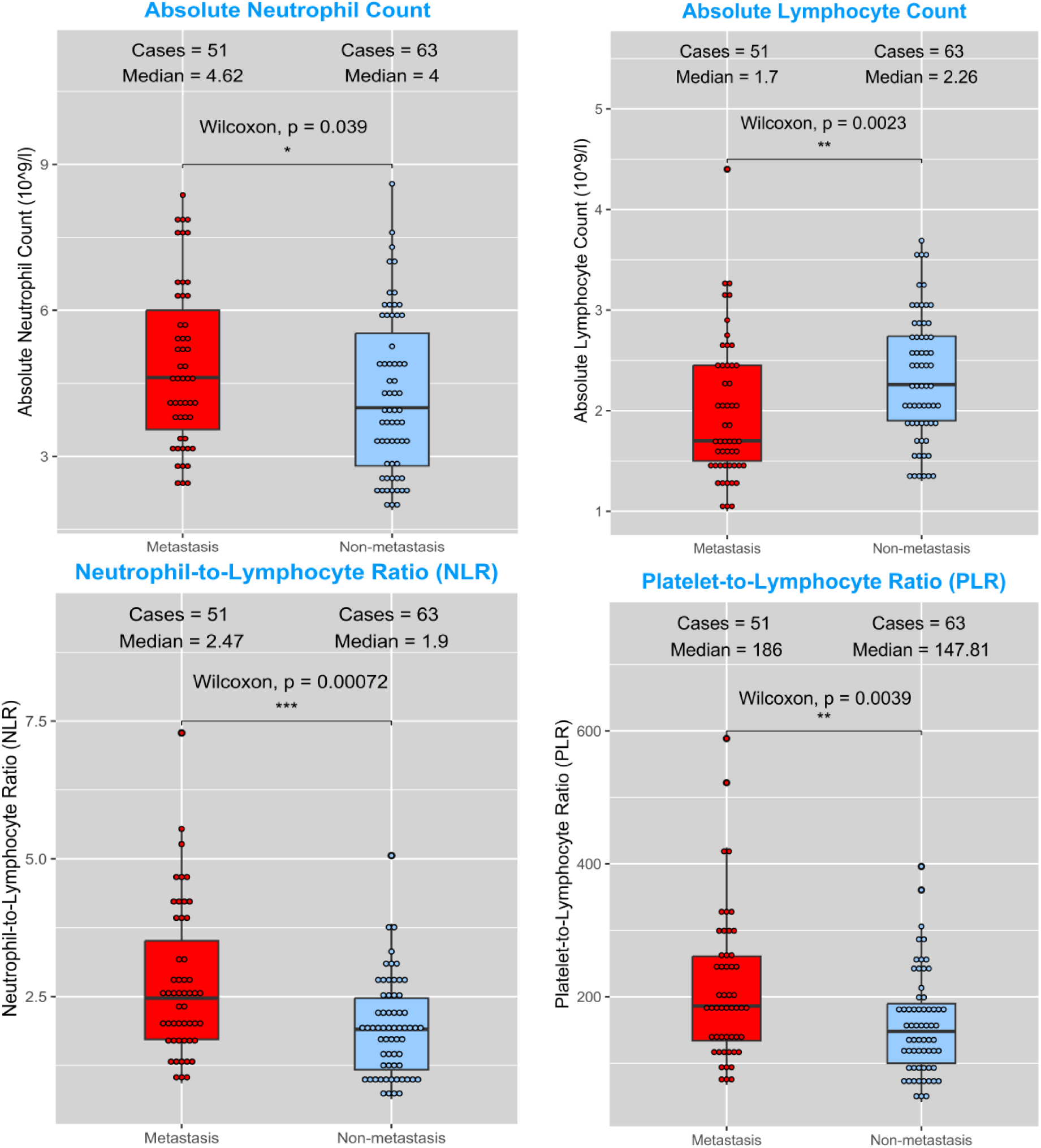
Comparison of absolute hematological counts and inflammatory ratios by metastatic status. Box plots illustrating significant differences in absolute neutrophil and lymphocyte counts, alongside the resulting elevations in NLR and PLR in patients with advanced metastatic gastric cancer.

Therefore, the neutrophil-to-lymphocyte ratio (NLR) was significantly elevated in the group of patients with distant metastases, median difference 0.56 (95% CI: 0.367–1.237), *p* = 0.00072 (Wilcoxon test). Similarly, a significant difference in the platelet-to-lymphocyte ratio (PLR) was observed between the two patient groups, median difference 38.19 (95% CI: 17.78–89.95), *p* = 0.0039 (Wilcoxon test) (**Figure 1**).

### Increased NLR and PLR correlated with disease progression

Our study next investigated the dynamic change of NLR and PLR across different stages. Our study here showed that the median NLR was 1.47 in stage I gradually rising to 1.84, stage II, 1.98 and stage III, and reached the highest value of 2.47 when distant metastasis was present (stage IV). It has been shown that the NLR is significantly different between stages II and IV (p = 0.00055, Wilcoxon test) and between stages III and IV (p = 0.031, Wilcoxon test), but no difference was observed in stage I and IV (**Figure 2**).

**Figure 2.**
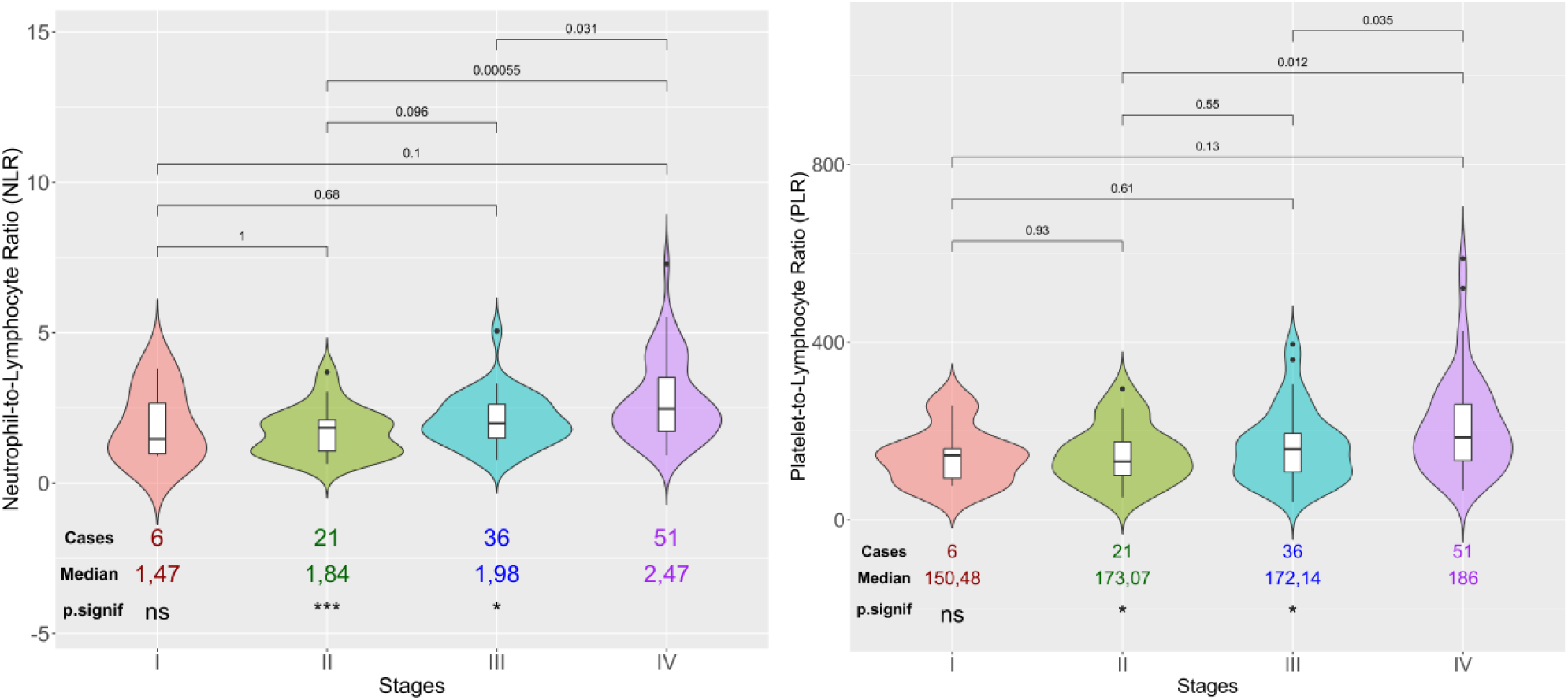
Progression of systemic inflammatory indices across pathological stages. The systemic inflammatory response, represented by NLR and PLR, demonstrates a stepwise increase as gastric cancer progresses through more advanced TNM stages.

Similarly, PLR displayed a stage-dependent increase. The median PLR was 150.48 in stage I, 173.07 in stage II, 172.14 in stage III, and 186.00 in stage IV. When comparing the PLR values between stage II and stage IV, as well as between stage III and stage IV, the results also indicated a statistical elevation (Wilcoxon test, p = 0.012 and p = 0.035, respectively) (**Figure 2**).

Our study also demonstrated that both NLR and PLR had a positive correlation with the progression of disease stages, with Spearman’s correlation coefficients of *r* = 0.33 and *r* = 0.27, respectively (*p* < 0.05), indicating that NLR and PLR tended to increase with disease advances. Additionally, NLR was positively correlated with PLR (*r* = 0.63). Furthermore, no correlation was observed between NLR or PLR and physiological factors, such as age or BMI, in gastric cancer patients in this study (**Figure 3**).

**Figure 3.**
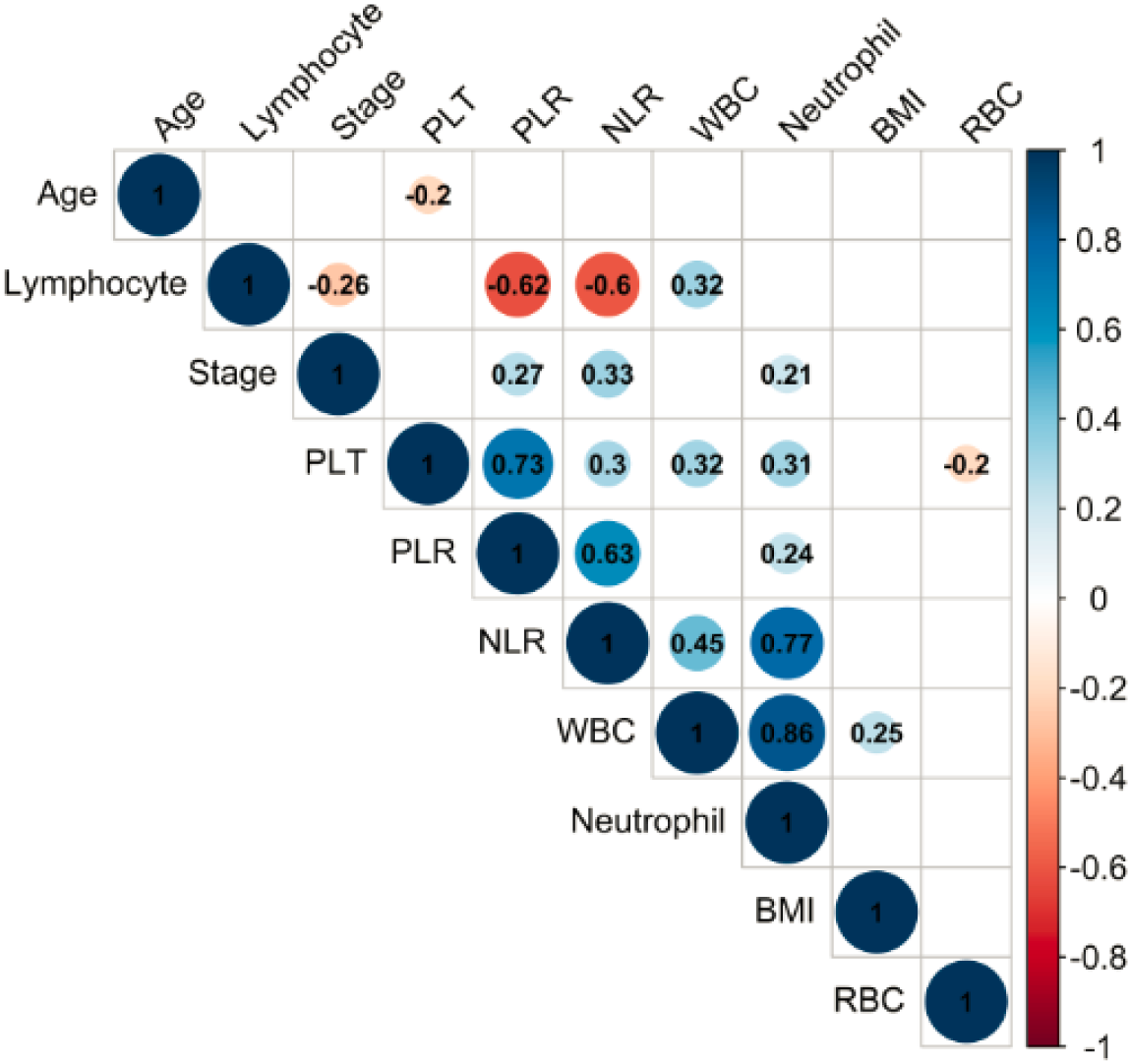
Correlation map of systemic inflammatory markers and clinicopathological features. Visualization of the interrelationships between NLR, PLR, disease stage, and key patient characteristics. Significant correlations are indicated by color intensity.

### Predictive value of the combined model integrating NLR, PLR, and clinical factors for gastric cancer metastasis

ROC curve analysis and the Youden index were used to determine the optimal cut-off values and predictive efficiency of NLR and PLR for GC with distant metastasis. The results indicated that, with a cut-off value of 2.0, NLR exhibited a sensitivity of 64.7% and specificity of 63.5% in predicting distant metastasis in GC (**Figure 4A**). For PLR, the optimal cutoff value was 181.5 with a sensitivity of 60.8% and a specificity of 73.0% (**Figure 4B**).

**Figure 4.**
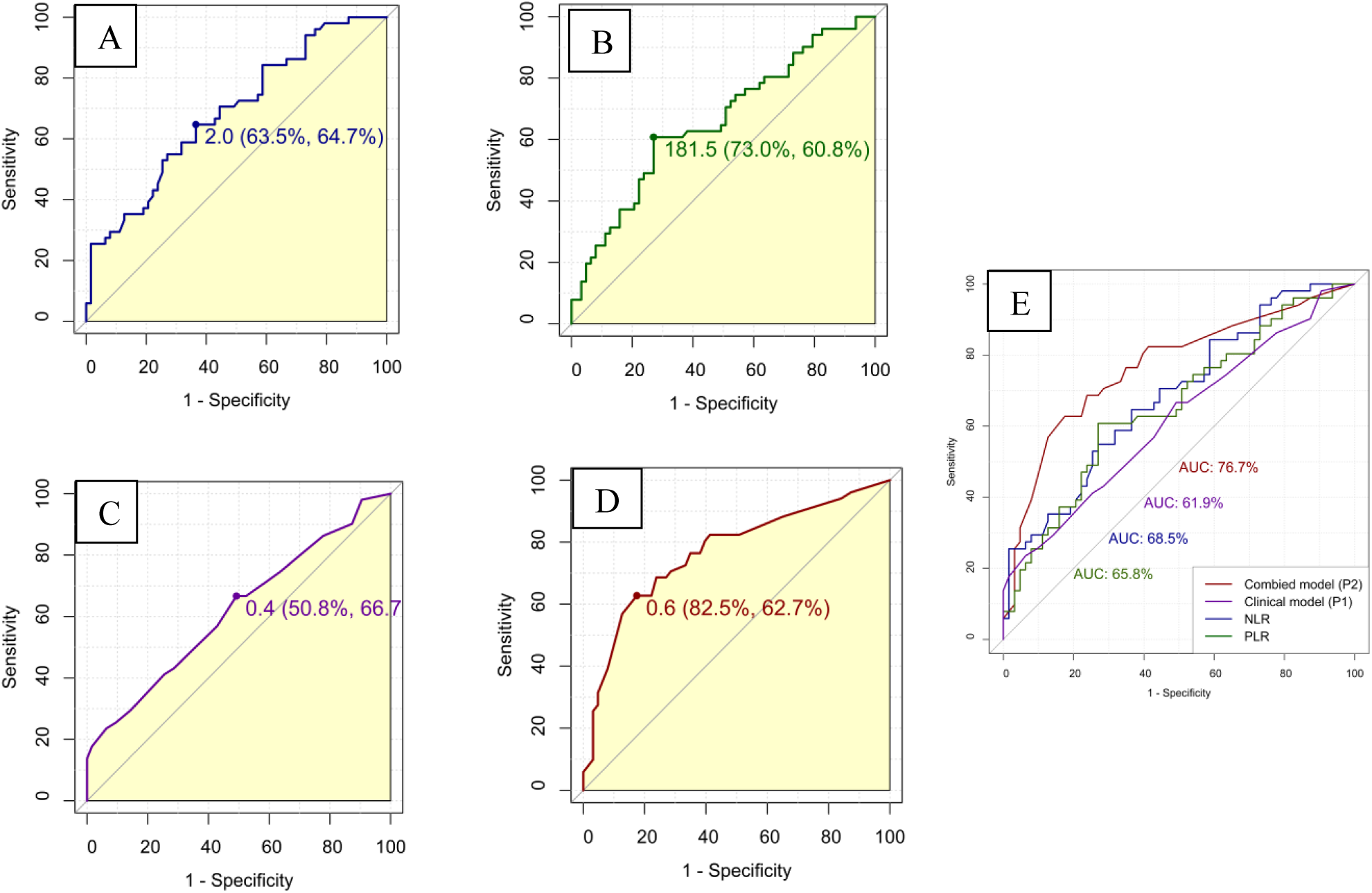
Receiver Operating Characteristic (ROC) analysis and model comparison. (A–B) ROC curves for individual inflammatory markers (NLR and PLR, respectively) with indicated optimal cut-off points. (C) Predictive performance of the baseline clinical model (Model P1). (D) Predictive performance of the integrated model combining clinical features with NLR and PLR (Model P2). (E) Direct comparison of AUC values between individual markers and the integrated models, demonstrating the superior performance of the combined approach.

The probability of predicting distant metastasis in GC was assessed using binary logistic regression for two models: one incorporating only clinical factors and another integrating both clinical factors with the NLR and PLR. The clinical factors analyzed in the regression analysis included age, sex, BMI, primary tumor location, and histological differentiation. The variables were categorized as follows: age (>60 vs. ≤60 years), sex (male vs. female), BMI (≥18.5 vs. <18.5 kg/m²), primary tumor location (multi-site vs. single-site), histological differentiation (poorly differentiated vs. moderately to well differentiated), NLR (≥2.0 vs. <2.0), and PLR (≥181.5 vs. <181.5).

The optimal cutoff value for the model incorporating only clinical factors (Model P1) was 0.4, with a sensitivity of 66.7% and a specificity of 50.8%. When NLR and PLR were combined with clinical factors (Model P2), the optimal cutoff value was determined to be 0.6, with a sensitivity of 62.7% and a specificity of 82.5% (**Figure 4C and 4D**).

The AUC for NLR in predicting distant metastasis in GC was 0.685 (95% CI: 0.587–0.782), while that for PLR was 0.658 (95% CI: 0.556–0.759). The AUC for the clinical factor-based model was 0.619 (95% CI: 0.515–0.723), whereas the combined model achieved an AUC of 0.766 (95% CI: 0.676–0.857) (**Figure 4E**).

The AUC of Model P1 (clinical factors only) was used as a reference to compare the predictive performance of NLR, PLR, and the combined model (Model P2). The results demonstrated that incorporating NLR and PLR into the clinical model significantly improved the predictive accuracy for distant metastasis (DeLong test, p = 0.00356) (**Table 2**).

**Table 2.** Diagnostic performance and predictive accuracy of inflammatory markers and clinical models for distant metastasis. Comparison of the area under the curve (AUC), optimal cut-off values (determined by the Youden index), sensitivity, specificity, and predictive values. Model P1 includes age (categorized as ≤60 vs. >60 years), sex (female vs. male), BMI (≥18.5 vs. <18.5 kg/m²), primary tumor location (single-site vs. multi-site), histological differentiation (poorly differentiated vs. moderately to well differentiated). Model P2 integrates these clinical features with NLR (<2.0 vs. ≥2.0) and PLR (<181.5 vs. ≥181.5). P values for AUC comparisons were calculated using DeLong’s test. Abbreviations: CI, confidence interval; Sen, sensitivity; Spe, specificity; PPV, positive predictive value; NPV, negative predictive value; Ref, reference.

|  | AUC | CI 95% | Cutoff | Sen | Spec | PPV | NPV | p-value |
| --- | --- | --- | --- | --- | --- | --- | --- | --- |
| **NLR** | 0.685 | 0.587 – 0.782 | 2.0 | 0.647 | 0.635 | 0.589 | 0.689 | 0.339 |
| **PLR** | 0.658 | 0.556 – 0.759 | 181.5 | 0.608 | 0.730 | 0.646 | 0.697 | 0.573 |
| **P2** | 0.766 | 0.676 – 0.857 | 0.59 | 0.627 | 0.825 | 0.744 | 0.732 | **0.00356** |
| **P1** | **0.619** | **0.515 – 0.723** | **0.41** | **0.667** | **0.508** | **0.523** | **0.653** | **Ref** |

In the combined model, univariate logistic regression analysis indicated that GC tumors with multiple locations (OR: 7.44; 95% CI: 1.84–50.0, *p* = 0.012), high NLR (cut-off: 2.0) (OR: 3.19; 95% CI: 1.49–7.00, *p* = 0.003), and high PLR (cut-off 181.5) (OR: 4.19; 95% CI: 1.93–9.44, *p* < 0.001) were associated with distant metastasis. However, when integrating these 3 significant variables in the multivariable regression analysis, only multiple tumor locations and PLR remained independent prognostic factors for the risk of distant metastasis, with ORs of 7.64 (95% CI: 1.71–54.9, *p* = 0.016) and 2.95 (95% CI: 1.17–7.65, *p* = 0.023), respectively (**Table 3**).

**Table 3.** Univariate and multivariate binary logistic regression analyses of risk factors associated with distant metastasis. Odds Ratios (OR) and 95% Confidence Intervals (CI) are presented. Only variables significant in univariate analysis were included in the final multivariate model.

| Characteristics | Univariate Analysis |  |  | Multivariate Analysis |  |  |
| --- | --- | --- | --- | --- | --- | --- |
|  | OR | 95% CI | p-value | OR | 95% CI | p value |
| **Age** |  |  |  |  |  |  |
| ≤ 60yrs | 1.00 | — |  | — | — | — |
| > 60yrs | 1.20 | 0.57–2.50 | 0.63 | — | — | — |
| **Sex** |  |  |  |  |  |  |
| Female | 1.00 | — |  | — | — | — |
| Male | 0.66 | 0.31–1.41 | 0.28 | — | — | — |
| **BMI** |  |  |  |  |  |  |
| **≥ 18.5** | 1.00 | — |  | — | — | — |
| **< 18.5** | 0.76 | 0.33–1.69 | 0.49 | — | — | — |
| **Histological grading** |  |  |  |  |  |  |
| **Intermediate/**<br>**Well differentiation** | 1.00 | — |  | — | — | — |
| **Poor differentiation** | 0.93 | 0.44–1.96 | 0.84 | — | — | — |
| **Tumor location** |  |  |  |  |  |  |
| **Single site** | 1.00 |  |  | 1.00 | — | — |
| **Multiple sites** | 7.44 | 1.84–50.0 | **0.012** | 7.64 | 1.71–54.9 | **0.016** |
| **NLR** |  |  |  |  |  |  |
| **Low risk (NLR < 2.0)** | 1.00 |  |  | 1.00 | — |  |
| **High risk (NLR ≥2.0)** | 3.19 | 1.49–7.00 | **0.003** | 1.92 | 0.75–4.88 | 0.2 |
| **PLR** |  |  |  |  |  |  |
| **Low risk (PLR <181.5)** | 1.00 |  |  | 1.00 | — |  |
| **High risk (PLR ≥181.5)** | 4.19 | 1.93–9.44 | **<0.001** | 2.95 | 1.17–7.65 | **0.023** |

## Discussion

### Predictive performance of NLR and PLR for distant metastasis in gastric cancer

Our study demonstrated that NLR and PLR were significantly elevated in patients with gastric cancer with distant metastasis, which is consistent with previous global studies.^[29–31]^ Likewise, our study agreed with previous report that increased PLR in GC patients was linked to a higher risk of advanced disease stage compared to those with lower PLR levels.^[32]^ Furthermore, we observed a progressive increase in NLR and PLR values, which correlated with disease stage advancement. The significant differences in NLR between stages II and IV, as well as between stages III and IV, underscore the potential utility of NLR in distinguishing locally advanced disease from metastatic gastric cancer. In contrast, the absence of a significant difference between stages I and IV suggests that NLR alone may have limited sensitivity in capturing early stage disease possibly due to the relatively preserved immune homeostasis in localized tumors or may also be partially attributable to the limited number of patients with stage I disease in our cohort (6 cases). Collectively, these results support the role of NLR as a marker of disease progression rather than early detection, highlighting its potential value in advanced-stage risk stratification and prognosis assessment.

In our study, we used the Youden index to determine the optimal NLR and PLR thresholds for predicting distant metastasis in a cohort of 114 patients with GC. The results indicated that both NLR and PLR possess moderate predictive value for distant metastasis in gastric cancer (GC) patients, with optimal cut-off values of 2.0 for NLR and 181.5 for PLR. The ROC analysis revealed that NLR had a sensitivity of 64.7% and specificity of 63.5%, while PLR showed slightly lower sensitivity (60.78%) but higher specificity (73.02%). These findings align with the recognized role of systemic inflammatory markers as accessible and cost-effective predictors of tumor progression and metastatic potential in solid tumors.

The threshold values of NLR and PLR for predicting the presence of distant metastasis in gastric cancer (GC) vary across studies. Abu-Shawer et al.^[29]^ identified an optimal NLR threshold of 3.9 for predicting distant metastasis in GC patients, whereas Zhang et al.^[30]^ proposed an NLR cutoff value of 2.91, with a sensitivity of 55% and specificity of 73.4%. Regarding PLR, in a meta-analysis by Zhang et al., including 49 studies comprising 51 cohorts, the authors noted that the PLR cutoff values ranged from 10.1 to 350, with a threshold of >150 being associated with poorer overall survival in GC patients.^[32]^

The observed variability in predictive NLR thresholds across existing literature may be attributed to heterogeneities in the study population characteristics, such as ethnicity, sex distribution, mean age, and disease stage proportions. Indeed, baseline NLR levels have been reported to vary with age within the same age group and across different ethnicities.^[33–35]^ Consequently, one future direction of for our study would be to include larger, multi-center cohorts to enhance the generalizability of these findings.

### Enhanced predictive accuracy through combined clinical and biomarker models

The AUC for NLR and PLR in predicting distant metastasis in GC were 0.685 and 0.658, respectively. Integrating NLR and PLR with established clinical factors (age, sex, BMI, primary tumor location, histological differentiation–categorized as binary groups) significantly improved the predictive accuracy for distant metastasis compared to clinical factors alone. The combined model (Model P2) achieved an AUC of 0.766, outperforming the clinical-only model (Model P1; AUC 0.619) and individual biomarkers. This enhancement underscores the additive value of systemic inflammatory markers in risk stratification frameworks, highlighting their potential utility in clinical decision-making to identify high-risk GC patients more reliably.

Numerous studies have investigated the prognostic and predictive value of NLR, PLR, or their combination in the survival outcomes of patients with GC^[36–38]^ However, the potential application of NLR and PLR in combination with clinical factors for predicting distant metastasis, a critical stage that requires well-planned treatment strategies, remains relatively underexplored. To the best of our knowledge, this is the first investigation on prognostic value of NLR and PLR in a Vietnamese gastric cancer patients cohort using a population-specific cutoff, thereby shaping a framework for future disease management but also facilitating methodological alignment with international studies.

### Independent Prognostic Factors for Distant Metastasis

Multivariable regression analysis identified multiple tumor locations and elevated PLR as independent prognostic factors for distant metastasis in this Vietnamese GC cohort. While high NLR was significant in univariate analysis (OR = 3.19; p = 0.003), it lost independent significance when adjusted for other variables (OR = 1.92; p = 0.2). This also showed that PLR remained an independent prognostic factor upon multivariate logistic regression (OR = 2.95; p = 0.023), suggesting that PLR may have a stronger or more direct association with metastatic risk in this population. The significance of multiple tumor locations further emphasizes the complexity of tumor biology influencing metastatic spread (OR = 7.64; p = 0.016).

Although NLR demonstrated superior discriminative power for distant metastasis in univariate analysis, its loss of statistical significance in the multivariable model–while PLR remained significant–likely stems from the moderate correlation between these two markers (r=0.63). This suggests a degree of multicollinearity and a substantial overlap in the physiological information they provide, which may have attenuated the independent predictive value of NLR. From a biological perspective, while NLR reflects general systemic inflammation, the independent significance of PLR may specifically highlight the critical role of platelet-mediated tumor angiogenesis and immune evasion—mechanisms that appear to be more direct drivers of distant spread in gastric cancer than the pathways represented by NLR alone.

### Clinical Implications, Limitations and Future Directions

The burden of GC in Vietnam closely parallels the global pattern, with most cases diagnosed at an advanced stage, leading to poor clinical outcomes. Alarmingly, the incidence of GC among younger individuals is rising rapidly, underscoring an urgent unmet need for earlier and more accurate risk stratification. However, robust data on readily-available, minimally invasive, cost-effective biomarkers with strong predictive value remain critically lacking in Vietnam where cancer patients generally face socio-economic burden and healthcare-related shortage. This study was designed to address this significant knowledge gap and represents the first and only Vietnamese dataset investigating this emerging clinical challenge.

These findings support the integration of NLR and PLR into routine clinical assessments to enhance early detection of metastatic risk, allowing for subsequent disease management strategies and optimal resource allocation. Hence, the available hematological parameters from routine complete blood count are potent tools for developing predictive models for clinical application. Use established cut-off values (NLR ≥ 2.0, PLR ≥ 181.5) to stratify patients into higher- and lower-risk categories for distant metastasis. Moreover, adopting a combined predictive model that integrates NLR, PLR and clinical features can improve accuracy in identifying patients at greatest risk and patients with elevated ratios should be considered for thorough screening, including more frequent imaging and clinical evaluations to detect early metastatic spread.

Despite the promising results, several limitations of this study warrant consideration. The single-center, retrospective design and the relatively modest sample size may restrict the generalizability of our findings. Future research should focus on validating these findings in larger cohorts to confirm the prognostic utility and refine cut-off values. Additionally, the correlation between NLR and PLR suggests that their predictive values are not entirely independent, leading to the attrition of NLR in our multivariable model. Moreover, further causal analyses should be employed to explore whether the prognostic impact of neutrophil-driven inflammation is fundamentally channeled through platelet-mediated pathways, thus discovering potential prevention strategies and treatment targets.

## Conclusion

In conclusion, our findings suggest that NLR and PLR increase with disease progression in patients with gastric cancer. Integrating these inflammatory indices with standard clinical features into a predictive model enhances the detection of distant metastasis, offering a cost-effective and accessible tool that requires no additional specialized equipment. Such a model holds substantial potential for the initial assessment and risk stratification of patients, particularly in clinical environments where advanced imaging may be constrained. Importantly, this study represents the first investigation in Vietnam to comprehensively evaluate the clinical relevance of these markers. These results lay a foundation for future multicenter, prospective studies to validate biomarker-based models, with the ultimate goal of optimizing management strategies for gastric cancer within resource-limited healthcare settings.

## Data Availability

The datasets generated and analyzed during the current study are available from the corresponding author upon reasonable request.

## Acknowledgements

We extend our deepest appreciation to the Board of Directors of the Breast, Gastroenterology, and Hepatology Department, as well as Ho Chi Minh City Oncology Hospital, for granting approval and providing essential support for this study.

## Declarations

### Ethics Approval and Consent to Participate

This study was conducted in accordance with the Declaration of Helsinki and received formal approval from the Ethics Committee of Ho Chi Minh City Oncology Hospital (Approval No. 292/BVUB-HĐĐĐ, dated April 26, 2023). As a retrospective chart review, the requirement for informed consent was waived. All patient data were anonymized and kept strictly confidential, used solely for the purposes of this research.

## Author Contributions

**Nhan Tran:** Conceptualization; Data curation; Formal analysis; Visualization; Writing – original draft; Writing – review & editing.

**Linh Tran Dang Ngoc:** Data curation; Investigation; Formal analysis; Writing – review & editing.

**Quy Nguyen Hoang:** Conceptualization; Methodology; Writing – review & editing; Supervision; Validation.

**Data Availability Statement:** The datasets generated and analyzed during the current study are available from the corresponding author upon reasonable request.

