## Supplementary material for "Predictive Value of Neutrophil-to-Lymphocyte and Platelet-to-Lymphocyte Ratios for Distant Metastasis in Vietnamese Gastric Cancer Patients": Table S1

### SUPPLEMENTAL FILE

**Table S1.** STROBE Statement—Checklist of items that should be included in reports of *cohort studies*

| | Item No | Recommendation | Page No |
|---|---|---|---|
| **Title and abstract** | 1 | (*a*) Indicate the study's design with a commonly used term in the title or the abstract | Retrospective cohort study as stated in the Abstract on page 1 and Methods on page 5 |
| | | (*b*) Provide in the abstract an informative and balanced summary of what was done and what was found | Provided in Abstract on page 1. |
| **Introduction** | | | |
| Background/rationale | 2 | Explain the scientific background and rationale for the investigation being reported | Included in the Introduction on pages 3. |
| Objectives | 3 | State specific objectives, including any prespecified hypotheses | Included in the Introduction on page 4. |
| **Methods** | | | |
| Study design | 4 | Present key elements of study design early in the paper | Included in the Methods on page 5. |
| Setting | 5 | Describe the setting, locations, and relevant dates, including periods of recruitment, exposure, follow-up, and data collection | Included in the Methods on pages 5. |
| Participants | 6 | (*a*) Give the eligibility criteria, and the sources and methods of selection of participants. Describe methods of follow-up | Included in the Methods on pages 5. |
| | | (*b*) For matched studies, give matching criteria and number of exposed and unexposed | Not applicable. |
| Variables | 7 | Clearly define all outcomes, exposures, predictors, potential confounders, and effect modifiers. Give diagnostic criteria, if applicable | Included in the Methods on pages 5, and potential confounders were further explained in Discussion on page 17. |

| | | | |
|---|---|---|---|
| Data sources/ measurement | 8* | For each variable of interest, give sources of data and details of methods of assessment (measurement). Describe comparability of assessment methods if there is more than one group | Included in the Methods on pages 5. |
| Bias | 9 | Describe any efforts to address potential sources of bias | Addressed in the limitations paragraph on Page 17 in the Discussion. |
| Study size | 10 | Explain how the study size was arrived at | Included in the Methods on pages 5 (The study size was determined based on the availability of eligible patients at our institution between January 2023 and June 2023. No formal sample size calculation was performed, as this was a retrospective cohort study. All patients who met the inclusion criteria during the study period were included in the analysis to maximize statistical power and representativeness). |
| Quantitative variables | 11 | Explain how quantitative variables were handled in the analyses. If applicable, describe which groupings were chosen and why | Included in the Methods on pages 5 and 6. |
| Statistical methods | 12 | (*a*) Describe all statistical methods, including those used to control for confounding | Included in the Methods on pages 5 and 6. |
| | | (*b*) Describe any methods used to examine subgroups and interactions | Included in the Methods on pages 6. |
| | | (*c*) Explain how missing data were addressed | Not applicable. Patients with missing data were excluded from the study. |
| | | (*d*) If applicable, explain how loss to follow-up was addressed | Not applicable. |
| | | (*e*) Describe any sensitivity analyses | Not applicable. |
| **Results** | | | |
| Participants | 13* | (a) Report numbers of individuals at each stage of study—eg numbers potentially eligible, examined for eligibility, confirmed eligible, included in the study, | Included in the Results on pages 6 and 7. |

| | | | |
|---|---|---|---|
| | | completing follow-up, and analysed | |
| | | (b) Give reasons for non-participation at each stage | Not applicable. |
| | | (c) Consider use of a flow diagram | Not required. |
| Descriptive data | 14* | (a) Give characteristics of study participants (eg demographic, clinical, social) and information on exposures and potential confounders | Included in the Results on page 6 and Table 1. |
| | | (b) Indicate number of participants with missing data for each variable of interest | Not applicable. Patients with missing data were excluded from the study. |
| | | (c) Summarise follow-up time (eg, average and total amount) | Not applicable. |
| Outcome data | 15* | Report numbers of outcome events or summary measures over time | Included in the Results on page 6, and summarized in Table 1. |
| Main results | 16 | (*a*) Give unadjusted estimates and, if applicable, confounder-adjusted estimates and their precision (eg, 95% confidence interval). Make clear which confounders were adjusted for and why they were included | Included in the Results on pages 5 and 6, and summarized in Tables 4 and 5. |
| | | (*b*) Report category boundaries when continuous variables were categorized | Included in the Results on pages 9 – 11, and summarized in Figure 1 and 2. |
| | | (*c*) If relevant, consider translating estimates of relative risk into absolute risk for a meaningful time period | Not applicable. |
| Other analyses | 17 | Report other analyses done—eg analyses of subgroups and interactions, and sensitivity analyses | Included in the Results on page 12 and Figure 4, and summarized in Table 2 and 3. |
| **Discussion** | | | |
| Key results | 18 | Summarise key results with reference to study objectives | Included in the Discussion on page 15, 16 and 17. |
| Limitations | 19 | Discuss limitations of the study, taking into account sources of potential bias or imprecision. Discuss both direction and magnitude of any potential bias | Addressed in the limitations paragraph on Page 17 in the Discussion. |
| Interpretation | 20 | Give a cautious overall interpretation of results considering objectives, | Included in the Discussion on pages 17. |

| | | | |
|---|---|---|---|
| | | limitations, multiplicity of analyses, results from similar studies, and other relevant evidence | |
| Generalisability | 21 | Discuss the generalisability (external validity) of the study results | Included in the Discussion on page 17. |
| **Other information** | | | |
| Funding | 22 | Give the source of funding and the role of the funders for the present study and, if applicable, for the original study on which the present article is based | Addressed in the Declarations on pages 18. |

*Give information separately for exposed and unexposed groups.
